# State of the evidence: a survey of global disparities in clinical trials

**DOI:** 10.1101/2020.10.08.20209353

**Authors:** Iain J Marshall, Veline L’Esperence, Rachel Marshall, James Thomas, Anna Noel-Storr, Frank Soboczenski, Benjamin Nye, Ani Nenkova, Byron C Wallace

**Affiliations:** School of Population Health and Environmental Sciences, King’s College London, London, UK; Cochrane Editorial and Methods Department, London, UK; EPPI-Centre, UCL, London, UK; Cochrane Dementia group, University of Oxford, Oxford, UK; Khoury College of Computer Sciences, Northeastern University, Boston, US; Computer and Information Science, University of Pennsylvania, Philadelphia, US

## Abstract

**Introduction:** Ideally, health conditions causing the greatest global disease burden should attract increased research attention. We conducted a comprehensive global study investigating the number of randomised controlled trials (RCTs) published on different health conditions, and how this compares with the global disease burden that they impose.

**Methods:** We use machine learning to monitor PubMed daily, and find and analyse RCT reports. We assessed RCTs investigating the leading causes of morbidity and mortality from the Global Burden of Disease study. Using regression models, we compared numbers of actual RCTs in different health conditions to numbers predicted from their global disease burden (Disability-Adjusted Life Years [DALYs]). We investigated whether RCT numbers differed for conditions disproportionately affecting countries with lower socio-economic development.

**Results:** We estimate 463,000 articles describing RCTs (95% prediction interval 439,000–485,000) were published from 1990 to July 2020. RCTs recruited a median of 72 participants (interquartile range 32–195). 82% of RCTs were conducted by researchers in the top fifth of countries by socio-economic development. As DALYs increased for a particular health condition by 10%, the number of RCTs in the same year increased by 5% (3.2%–6.9%), but the association was weak (adjusted R^2^=0.13). Conditions disproportionately affecting countries with lower socio-economic development, including respiratory infections and tuberculosis (7 thousand RCTs below predicted) and enteric infections (10 thousand RCTs below predicted), appear relatively under-researched for their disease burden. Each 10% shift in DALYs towards countries with low and middle socio-economic development was associated with a 4% reduction in RCTs (3.7%–4.9%). These disparities have not changed substantially over time.

**Conclusion:** Research priorities are not well optimized to reduce the global burden of disease. Most RCTs are produced by highly developed countries, and the health needs of these countries have been, on average, favoured.

**Key questions:** *What is already known?:*

- Prior studies have manually investigated the relationship between published research in different health conditions and the global burden of disease that they impose.
- However, these analyses have been mostly limited to estimates of research funding from national funders, or smaller scale analysis of older publication records.
- These studies have highlighted disparities in research relative to burden, but they are not sufficient to enable global targeting of research to optimise improvements in disease burden.

*What are the new findings?:*

- We automatically process all of PubMed, allowing us to conduct a continually updated, comprehensive analysis of published reports of RCTs, including the number of participants per RCT and the health conditions studied.
- We found that considerable disparities exist between the relative volume of evidence on some conditions and the global burden of disease that they impose, as calculated by the Global Burden of Disease study.
- Further, our analysis suggests that there exists a smaller amount of evidence for conditions that impose a comparatively large burden of disease in lower-income countries.

*What do the new findings imply?:*

- Looking at numbers of RCTs published, and the numbers of participants in these trials, it seems that research priorities are not optimized to reduce the global burden of disease, and that research for conditions affecting higher-income countries has, on average, been favoured.
- The findings from this study could help research funders to focus research investment in areas where the largest reductions in disease burden could be made.

## Introduction

The Global Burden of Disease (GBD) study has estimated the leading causes of death and disability worldwide.[1] Health inequalities have been defined as differences in health which are avoidable, remediable, and considered unjust.[2] A key strategy to reduce death and morbidity is to conduct randomized controlled trials (RCTs) to determine the effects of interventions for improving health.[3] However, RCTs are expensive and time-consuming, and there is necessarily a limit on the number and size of these types of study that can be carried out. There are very many factors which could reasonably affect which health problems are researched, not limited to: the existence of highly effective treatments for certain conditions (where poor implementation or availability is the problem); national governments prioritising conditions which affect their own country’s population; and the unpredictable nature of scientific breakthroughs in therapy. Pharmaceutical companies are likely to develop drugs which have the potential to be sold at profit; this can lead to neglect of conditions (e.g. infectious diseases) which disproportionately affect low income countries.[4] Nonetheless, a rational approach to funding research — reduce health inequalities and maximise global benefit — would take account of the global disease burden.

Studies from the US and the UK have reported that disparities exist in national health research funding for certain conditions. Some conditions, such as diabetes and cancer, have attracted a comparatively generous amount of research funding in these countries relative to their national and global burden, whereas other conditions, such as stroke and respiratory diseases, have been relatively neglected.[5–8] In a later study, Evans and colleagues mapped the volume of research (RCTs and systematic reviews) published in 2005 to conditions studied in the Global Burden of Disease (GBD) study. They concluded that global Disability Adjusted Life Years (DALYs; a measure that incorporates the quality of lived years with respect to physical and mental functioning criteria) in 2004 did not explain *any* of the topic distribution of research articles published in the following year.[9] By contrast, Atal and colleagues examined 117,000 new registrations for clinical trials globally from 2006–2015, and reported that trials registered were proportionate to disease burden in high-income countries, but not in non-high income countries.[10]

To our knowledge, there has been no comprehensive, global study comparing published research (both in terms of trial and participant numbers) with the relative burden of each condition. We address this gap by evaluating the total volume of evidence (numbers of RCTs and trial participants) published for clinical conditions via an automated analysis of all research articles indexed in PubMed (http://pubmed.ncbi.nlm.nih.gov) from 1990 onwards. Our system uses natural language processing to automatically identify text within articles that describes the conditions being studied, and maps these to GBD-defined categories. This permits comparison between the global disease burden and the amount of published evidence that exists for each condition, and could help prioritise research funding to relatively understudied conditions with high burden. Uniquely, as our method is fully automatic, we are able to analyse articles at large scale (classification of 100’s of thousands of documents), and with continuous, live updating — neither of which have been feasible so far with manual methods.

## Methods

### Overview

We developed a machine learning system which maintains a live database of annotated RCT reports, named Trialstreamer. We have described the computational methods and accuracy of the system components in detail elsewhere,[11] and summarize the key points relevant to the current study below.

### Monitoring PubMed for RCTs in humans

We first downloaded the full PubMed database (annual baseline data, from 11th December 2018) and then subsequently monitored and added all newly published articles daily. This comprises >30 million articles. We automatically identify the subset of articles that describe RCTs using a machine learning system, which operates over the text of the title and abstract, and the *Publication Type* database index.[12] Briefly, we ensemble a Support Vector Machine (SVM)[13] and a Convolutional Neural Network (CNN).[14] These machine learning models are *trained*^1^ on 280,000 abstracts manually labelled as being RCTs or not by Cochrane Crowd,^2^ a collaborative citizen science project. We do not rely on the Publication Type index alone, as we have previously found it to miss a substantial proportion of the 5–7 most recent years of articles[11] (due to delay in manual indexing after publication). We next removed any RCTs that were not conducted in humans (e.g., animal or agricultural studies) using an SVM model. This model was trained using labelled abstracts from PubMed (labels derived as a function of whether the MeSH^3^ term ‘Humans’ had been applied or not).

### Automatic information extraction

To characterise the trial participants and outcomes, we first automatically extract ‘snippets’ of text (typically a few consecutive words within the abstract) describing these components. For this, we use a *sequence tagging* machine learning model (namely a Long Short-Term Memory network[15] followed by a conditional random field,[16] usually abbreviated to LSTM-CRF[17]).[18] This model was trained using a publicly available dataset called EBM-NLP, which comprises 5,000 RCT abstracts in which text spans relevant to the respective PICO characteristics (participants, interventions, comparators, and outcomes) were manually annotated.

These text descriptions are then standardised to MeSH terms. This allows us to infer, for example, that ‘myocardial infarction’ and ‘heart attack’ refer to the same condition, and that both are subtypes of cardiovascular disease. To achieve this step, we use an approach similar to that described by Demner-Fushman and colleagues, in their *MetaMap* software.[19] This approach involves generating a large database of text synonyms for MeSH terms, done by finding alternative descriptions for each term in linked vocabularies. These synonyms are then sought in the extracted population and outcomes text spans.

### Linking RCTs with data on global disease burden

In this study, we use the 22 level 2 categories defined by the GBD collaboration. These categories describe broad disease areas such as “Cardiovascular diseases”, and “Mental disorders” (the full list is provided in Table 1). The GBD collaboration makes available International Classification of Diseases (ICD-10) codes which define the scope of each category. We automatically link these ICD-10 codes to the equivalent MeSH terms using a tool and dataset named *Metathesaurus*, from the Unified Medical Language System (UMLS).[20] In brief, Metathesaurus provides a database of synonyms between different health science vocabularies, and thus lists of synonymous terms between ICD-10 and MeSH can be retrieved. One clinical author (IJM) manually checked all of the automatically generated mappings from MeSH to Global Burden of Disease category (both removing any errors from the automatic mapping, and adding links where they had been missed). This resulted in a final mapping of 5,256 parent MeSH terms to one of the 22 GBD categories.

**Table 1:** RCTs in PubMed with estimates of trial population sizes, and comparison with measures of global disease burden. DALY=Disability-adjusted life year.

| disease category | Total RCTs, thousands, (95% prediction interval) [rank] | RCTs with the condition as a *population* thousands, (95% prediction interval) [rank] | RCTs with the condition as an *outcome* thousands, (95% prediction interval) [rank] | DALYs (Disability-Adjusted Life Years), thousands [rank] | Death), thousands [rank] | YLDs (Years Lived with Disability), thousands [rank] | YLLs (Years of Life Lost, thousands [rank] |
| --- | --- | --- | --- | --- | --- | --- | --- |
| Mental disorders | 55 (49–61) [1] | 28 (25–31) [3] | 40 (36–44) [1] | 2,900 [11] | <0.1 [21] | 2,900 [2] | 0.4 [21] |
| Cardiovascular diseases | 54 (49–60) [2] | 37 (33–41) [1] | 31 (28–34) [2] | 8,700 [1] | 410 [1] | 710 [11] | 8,000 [1] |
| Other non-communicable diseases | 39 (35–43) [3] | 24 (21–26) [5] | 20 (18–22) [4] | 3,500 [6] | 33 [15] | 1,300 [6] | 2,200 [9] |
| Neurological disorders | 39 (35–43) [4] | 26 (23–28) [4] | 20 (18–23) [3] | 2,500 [12] | 58 [7] | 1,700 [3] | 820 [17] |
| Neoplasms | 31 (28–34) [5] | 29 (26–31) [2] | 6 (6–7) [12] | 5,400 [4] | 210 [2] | 150 [20] | 5,200 [4] |
| Maternal and neonatal disorders | 24 (22–26) [6] | 19 (17–21) [6] | 13 (11–14) [5] | 7,200 [2] | 77 [5] | 630 [13] | 6,500 [2] |
| Diabetes and kidney diseases | 21 (19–23) [7] | 18 (17–20) [7] | 5 (4–5) [13] | 2,200 [16] | 52 [9] | 900 [9] | 1,300 [15] |
| Unintentional injuries | 18 (16–20) [8] | 11 (10–12) [11] | 9 (8–10) [8] | 3,300 [7] | 51 [10] | 800 [10] | 2,500 [7] |
| Musculoskeletal disorders | 18 (16–20) [9] | 13 (12–15) [8] | 7 (6–8) [10] | 3,100 [8] | 2 [19] | 3,000 [1] | 65 [19] |
| Digestive diseases | 17 (15–19) [10] | 13 (12–14) [9] | 7 (6–8) [11] | 2,100 [18] | 56 [8] | 440 [14] | 1,700 [14] |
| Substance use disorders | 17 (15–19) [11] | 8 (7–8) [14] | 12 (11–13) [6] | 1,000 [22] | 8 [18] | 710 [12] | 310 [18] |
| Skin and subcutaneous diseases | 16 (15–18) [12] | 8 (7–9) [13] | 12 (11–13) [7] | 1,000 [21] | 2 [20] | 990 [7] | 55 [20] |
| Chronic respiratory diseases | 14 (13–16) [13] | 13 (11–14) [10] | 4 (4–5) [15] | 2,900 [10] | 99 [4] | 990 [8] | 1,900 [12] |
| Sense organ diseases | 14 (13–16) [14] | 8 (7–9) [12] | 8 (7–9) [9] | 1,400 [20] | n/a | 1,400 [4] | n/a |
| HIV/AIDS and sexually transmitted infections | 8 (7–9) [15] | 7 (6–8) [15] | 3 (3–4) [17] | 2,500 [13] | 41 [11] | 130 [21] | 2,300 [8] |
| Respiratory infections and tuberculosis | 8 (7–8) [16] | 4 (3–4) [17] | 5 (4–5) [14] | 6,500 [3] | 120 [3] | 290 [17] | 6,200 [3] |
| Other infectious diseases | 7 (6–8) [17] | 5 (4–5) [16] | 3 (2–3) [18] | 3,000 [9] | 39 [12] | 110 [22] | 2,900 [6] |
| Nutritional deficiencies | 5 (4–6) [18] | 2 (1–2) [19] | 4 (3–4) [16] | 2,100 [17] | 12 [17] | 1,300 [5] | 840 [16] |
| Neglected tropical diseases and malaria | 3 (3–4) [19] | 2 (2–3) [18] | 1 (1–1) [19] | 2,300 [14] | 27 [16] | 410 [15] | 1,900 [10] |
| Self-harm and interpersonal violence | 2 (1–2) [20] | 0.8 (0.7–0.9) [20] | 1.0 (0.9–1) [20] | 2,000 [19] | 37 [14] | 180 [19] | 1,900 [13] |
| Enteric infections | 1 (1–2) [21] | 0.6 (0.5–0.7) [21] | 0.8 (0.8–0.9) [21] | 4,100 [5] | 64 [6] | 240 [18] | 3,800 [5] |
| Transport injuries | 0.4 (0.4–0.5) [22] | 0.2 (0.2–0.2) [22] | 0.2 (0.2–0.2) [22] | 2,200 [15] | 38 [13] | 300 [16] | 1,900 [11] |

**Table 2:** Difference between measured number of RCTs, and number predicted based on global disease burden (negative numbers=fewer RCTs were published than predicted)

| Disease category | Total<br>[1990–<br>2017] | 1990–<br>1993 | 1994–<br>1997 | 1998–<br>2001 | 2002–<br>2005 | 2006–<br>2009 | 2010–<br>2013 | 2014–<br>2017 |
| --- | --- | --- | --- | --- | --- | --- | --- | --- |
| Cardiovascular diseases | +34,000 | +2,400 | +3,000 | +3,600 | +4,400 | +5,100 | +6,700 | +8,300 |
| Chronic respiratory diseases | +4,200 | +370 | +500 | +670 | +720 | +710 | +620 | +630 |
| Diabetes and kidney diseases | +10,000 | +490 | +710 | +800 | +1,200 | +1,600 | +2,400 | +3,400 |
| Digestive diseases | +7,700 | +590 | +850 | +920 | +1,100 | +1,100 | +1,400 | +1,700 |
| Enteric infections | -9,700 | -950 | -1,200 | -1,200 | -1,400 | -1,500 | -1,700 | -1,800 |
| HIV/AIDS and sexually transmitted infections | -1,100 | -120 | -180 | -190 | -400 | -280 | -51 | +100 |
| Maternal and neonatal disorders | +6,400 | +120 | +410 | +440 | +530 | +670 | +1,500 | +2,700 |
| Mental disorders | +38,000 | +1,100 | +1,900 | +2,600 | +4,100 | +6,100 | +9,200 | +13,000 |
| Musculoskeletal disorders | +5,900 | +50 | +130 | +360 | +780 | +1,000 | +1,600 | +2,000 |
| Neglected tropical diseases and malaria | -5,400 | -470 | -570 | -650 | -790 | -880 | -990 | -1,100 |
| Neoplasms | +14,000 | +980 | +1,300 | +1,200 | +1,700 | +2,200 | +3,000 | +4,000 |
| Neurological disorders | +25,000 | +1,100 | +1,700 | +2,100 | +2,800 | +3,800 | +5,600 | +7,800 |
| Nutritional deficiencies | -3,300 | -380 | -450 | -460 | -410 | -520 | -560 | -530 |
| Other infectious diseases | -2,500 | -390 | -400 | -340 | -350 | -260 | -350 | -410 |
| Other non-communicable diseases | +24,000 | +890 | +1,400 | +1,800 | +2,800 | +3,900 | +6,100 | +7,400 |
| Respiratory infections and tuberculosis | -7,000 | -830 | -980 | -970 | -1,100 | -1,200 | -1,100 | -1,000 |
| Self-harm and interpersonal violence | -6,800 | -620 | -840 | -890 | -920 | -1,000 | -1,200 | -1,300 |
| Sense organ diseases | +6,500 | +470 | +630 | +800 | +780 | +1,000 | +1,300 | +1,500 |
| Skin and subcutaneous diseases | +9,300 | +660 | +780 | +980 | +1,200 | +1,500 | +1,900 | +2,300 |
| Substance use disorders | +9,800 | +370 | +750 | +870 | +1,200 | +1,700 | +2,200 | +2,700 |
| Transport injuries | -8,000 | -670 | -840 | -940 | -1,100 | -1,200 | -1,500 | -1,700 |
| Unintentional injuries | +6,300 | +91 | +200 | +310 | +620 | +840 | +1,600 | +2,600 |

### Descriptive statistics

We report descriptive statistics for RCT counts and participant numbers. We use the country of the first author’s affiliation as a proxy for the country organising or funding the study. In order to capture trials addressing prevention, we also consider RCTs as relevant to the GBD category where rates of the health condition in question are identified as an *outcome*, even where participants did not have the condition at trial onset. For example, trials that recruited people with diabetes but measured the effects of an intervention on subsequent coronary heart disease were considered as relevant to both the ‘diabetes’ and ‘cardiovascular diseases’ categories. We also present separate estimates based on this distinction, that is, separating instances where a given GBD condition was found in the trial population (‘treatment’ trials) from those where it was inferred to describe an outcome (‘prevention’ trials).

### Estimating uncertainty due to prediction error

We randomly sampled articles (titles and abstracts) from the finalised dataset, and two authors independently manually labelled these with respect to study design (RCT in humans *vs*. not; 500 abstracts, done by IJM and BCW), Global Burden of Disease category (250 abstracts, done by IJM and VL), and number of participants randomized (500 abstracts, done by IJM and BCW). Authors then met to resolve discrepancies by consensus. We then used the bootstrap method to estimate 95% prediction intervals for final counts, using the method described by Fox and colleagues.[21,22] In short, we use our manually labeled data to calculate 10,000 bootstrap estimates of the precision and recall of each labeling task.[23] These precision and recall estimates are then used to adjust the automated results from the full dataset (generating 10,000 adjusted count estimates for each classification task), which can be used to construct a 95% prediction interval. To evaluate uncertainty around extracted sample sizes we use a similar approach, except we used estimates of the absolute error taken from the manually labeled abstracts. We then simulated the effect of this error on the full dataset using the bootstrap method with 10,000 iterations. These intervals permit assessment of the degree to which model misclassification and bias has affected the final results.

### Assessing associations between disease burden and published RCTs

To examine associations between global disease burden and volume of evidence (both numbers of RCTs and participants), we used a series of linear regression models with log-transformed global DALYs as the predictor, and the log-transformed numbers of global RCTs as the dependent variable. We evaluate annual data on DALYs for each clinical condition for the full period available from the Global Burden of Disease study (1990–2017), and adjust for trends over time by incorporating the year as a categorical predictive variable. A detailed description alongside statistical code and data for these analyses is available at the Open Science Framework (DOI: <u>10.17605/osf.io/3db76</u>).

In our first regression analysis, we examined the association of global DALYs on RCT publications in the same year. We explored incorporating the year (as categorical variable) both as a fixed and a random effect, evaluating using the Hausman test. We compare observed RCT counts for each GBD condition with those predicted by the model.

In our second regression analysis, we examine the effect of lag (given that RCTs will publish reports some time after measures of disease burden are known). Here we use finite length distributed lag models, and evaluate the association of global DALYs with up to 1, 2, 3, 4, and 5 years lag on published RCTs in the index year. We compare models using the adjusted R^2^ statistic, the Akaike information criterion (AIC), and Bayesian information criterion (BIC).

In our third regression analysis, we add a variable to account for whether a particular condition tends to impose a disproportionate burden in high-income, rather than low and middle-income countries.

We examine associations between socioeconomic status and evidence volume, using the Socio-Demographic Index (SDI) of the country from the Global Burden of Disease study. As a metric of disparity in disease burden, we used the ratio of DALYs occurring in high SDI countries (the top fifth) to DALYs occurring in low and medium SDI countries (i.e., the bottom four fifths).

Finally, we conducted a sensitivity analysis aiming to discover whether our results are sensitive to ‘salami slicing’ (the practice of publishing results from a single trial across multiple publications, which could lead us to overestimate the evidence base). We first estimated the mean number of publications per RCT for each GBD condition, utilising the subset of articles which contained a unique trial registry identifier. Then we use these values to discount our estimate of RCT numbers for each GBD condition. We repeated our regression analyses with these discounted RCT numbers.

## Results

We analysed records of 31,367,011 articles indexed in PubMed on July 27th 2020. We estimate that 459,000 (1.5%) of these articles were reports of RCTs conducted in humans (95% prediction interval 435,000 to 480,000). We show how these trials are distributed globally (by location of the first author) in Figure 1. The number of RCTs published for each of the 22 ‘level 2’ disease categories from the 2017 Global Burden of Disease study is presented in Table 1 and Figures 2 and 3.

**FIGURE 1:**
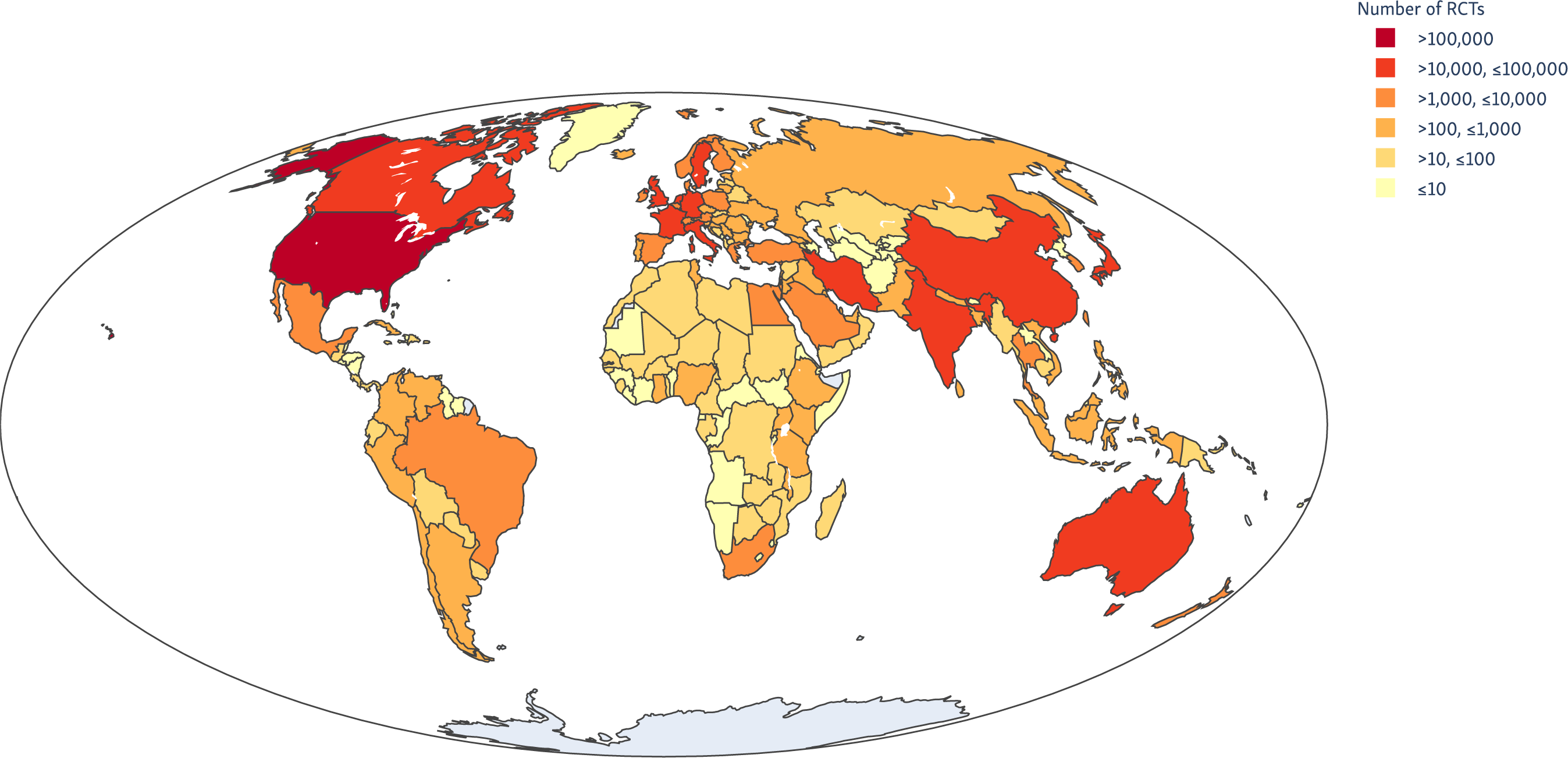
Global clinical trial publications by first author location, 1990–2017.

**FIGURE 2:**
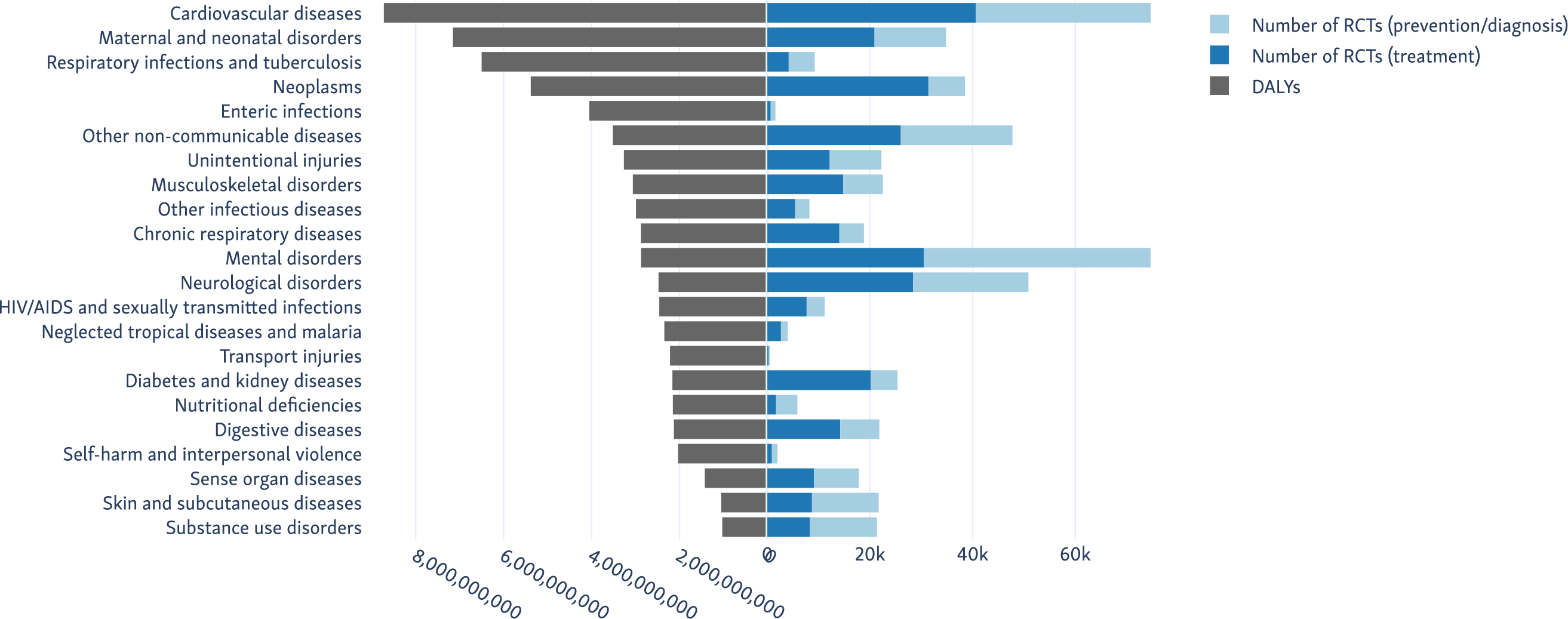
Disease categories causing the highest global burden (by DALY), with the number of RCTs published for each).

**FIGURE 3:**
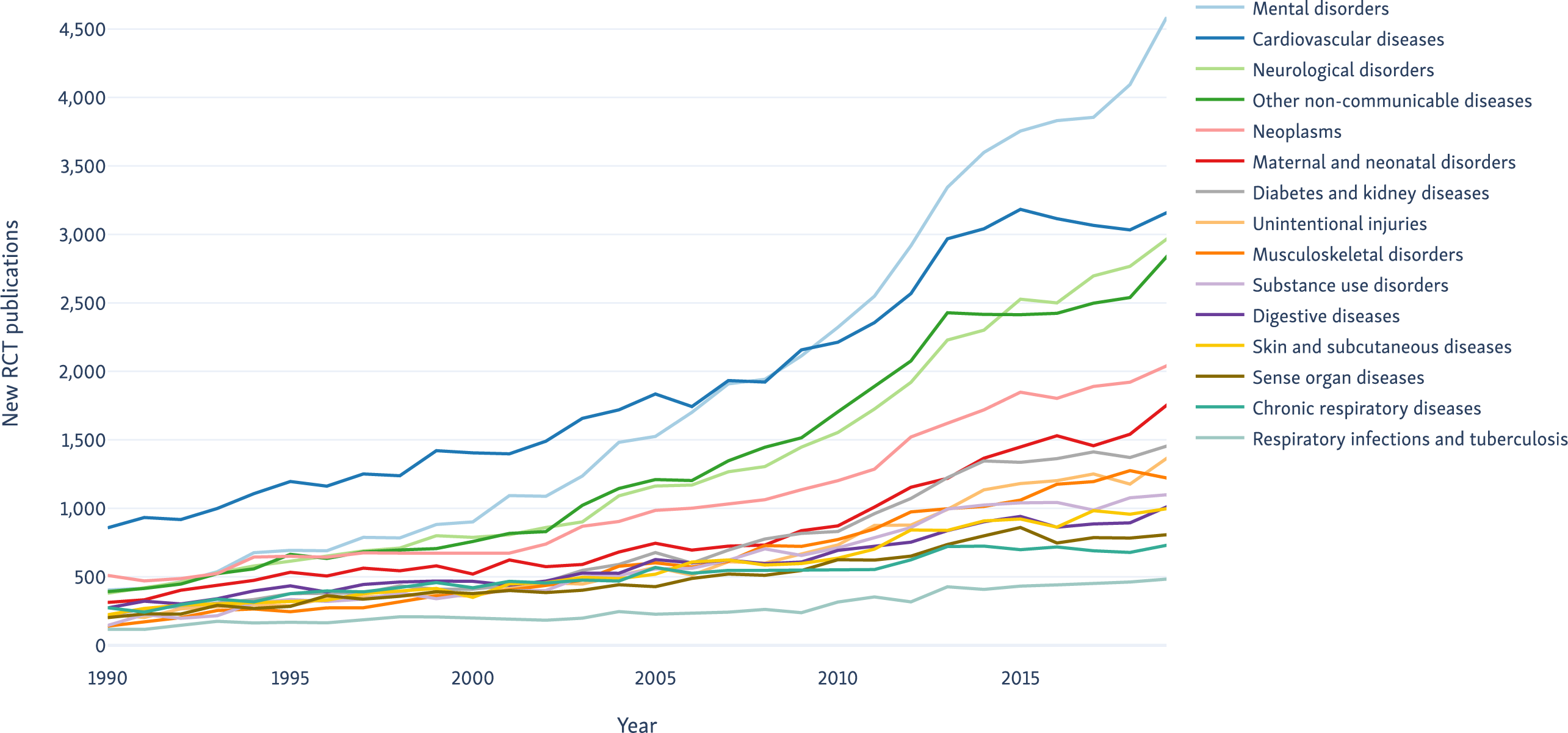
Trends over time in RCT publications of the top fifteen categories by publication rates from 1990 to 2019.

Mental disorders (ranked 2nd in terms of total DALYs) had the largest number of RCTs with 50,000, followed by Cardiovascular diseases (ranked 1st) with 49,000. We separate the results from trials where the clinical condition occurred in the trial population (generally trials of *treatment* strategies for), and where the condition was a trial outcome (typically trials looking at *prevention or diagnosis* of the condition) in Table 1 and Figure 2. In all cases except for Mental disorders, the numbers of trials and participants in the ‘treatment’ trials exceeded those in the ‘prevention’ trials.

Overall, we estimate the median number of participants randomized per RCT was 72, with interquartile range (IQR) 32 to 195. We show how the distribution of RCT sample sizes varies for each of the GBD categories in Figure 4.

**FIGURE 4:**
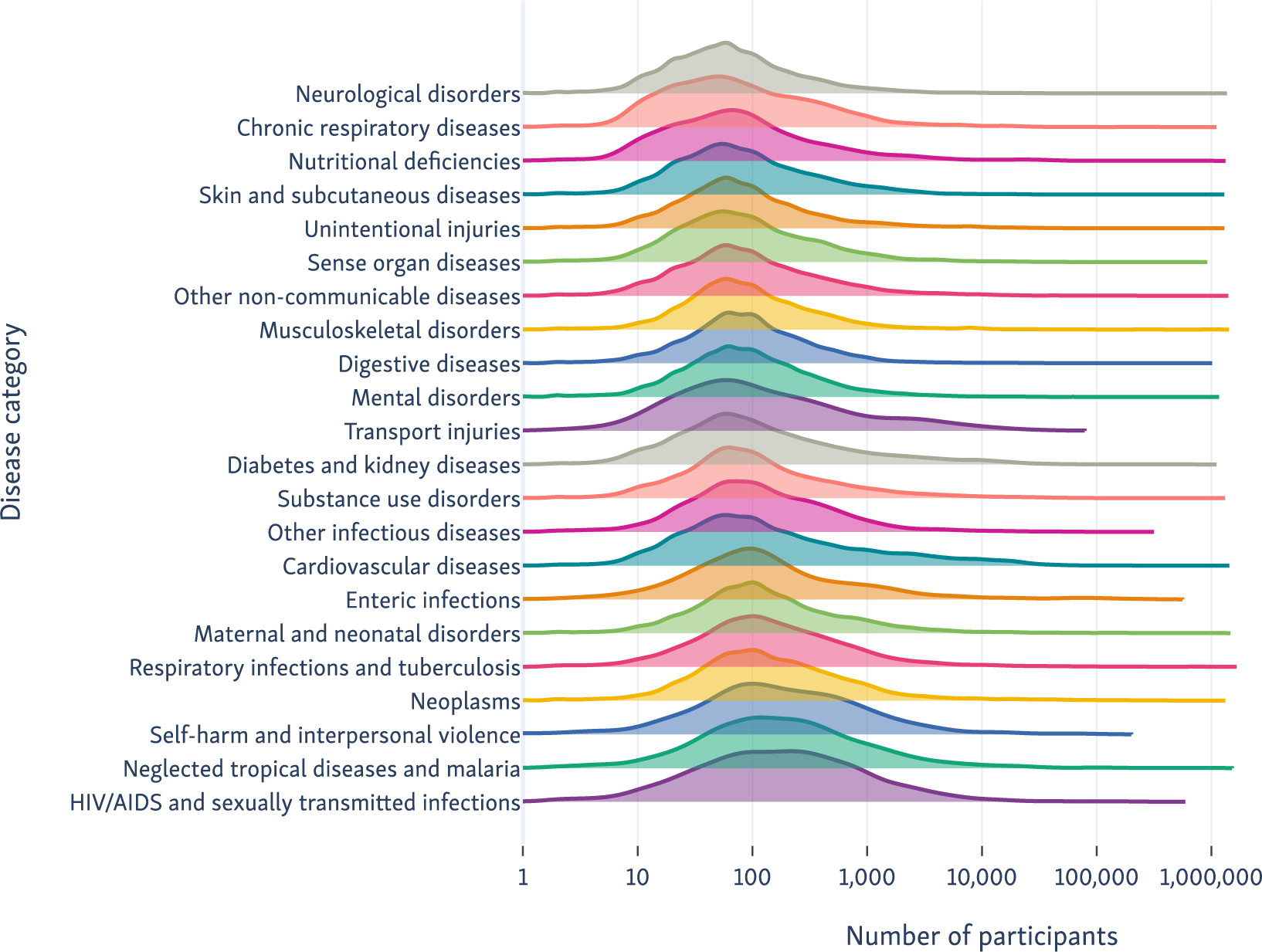
Number of participants recruited to RCTs pertaining to each of the Global Burden of Disease categories.

**FIGURE 5:**
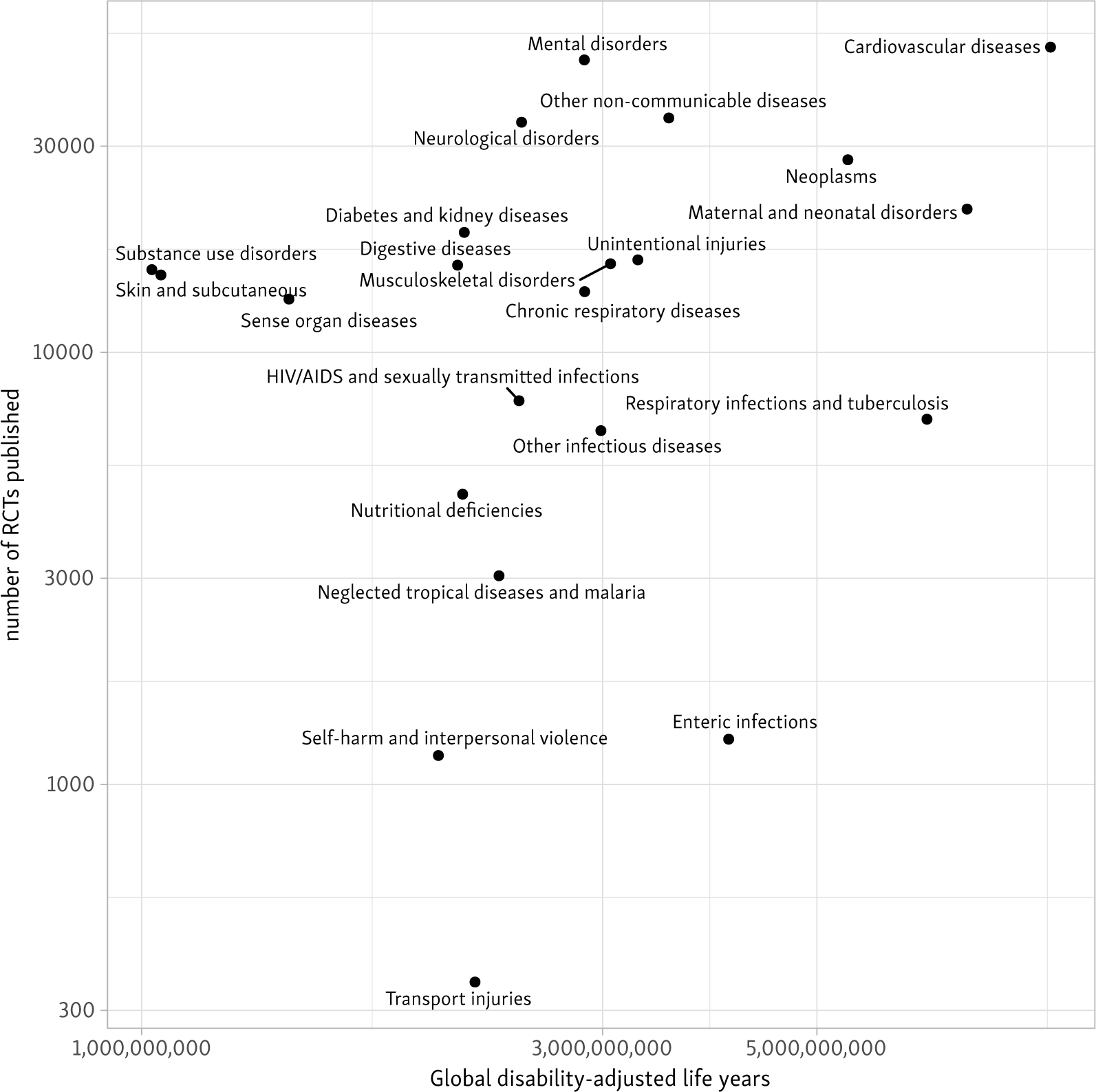
Total RCTs published globally from 1990–2017 against disability-adjusted life years in the same period.

In Figure 5 we scatter the volume of evidence estimated for conditions against the global burden that they impose (totals in period 1990–2017) for the 22 ‘level 2’ Global Burden of Disease categories.

Our primary analysis found that, on average, increasing DALYs were associated with a statistically significant increase in RCT publication in the same year, but that variance was not well explained by the model (β=0.52 for log DALYs, 95% confidence interval 0.33 to 0.70; R^2^=0.13). This translates to an average increase in the number of RCTs published of 5% (3.2% to 6.9%) for each 10% increase in DALYs in the same year. We report differences between the expected versus observed number of RCTs and trial participants based on this model in Table 2.

We found that models incorporating between 3–5 years of lag had improved fit compared with the equivalent model with no lag effects (demonstrated by reduction in Akaike information criterion [AIC], Bayesian information criterion [BIC], and increase in adjusted R^2^). The model incorporating lag up to 3 years found an average 7.2% increase in published RCTs for every 10% increase in DALYs (adjusted R^2^=0.29). The models with lag up to 4, and up to 5 years gave near identical estimates and fit (full analysis results provided in the statistical appendix).

We provide full results for our sensitivity analysis (where we adjust for the differential impact of RCTs producing multiple publications in each disease in our statistical appendix (https://osf.io/zf97n/). We found that RCTs with a registry identifier produced a mean of 1.3 articles, but that there was variation between GBD categories (from 1.2 articles/RCT in nutritional deficiencies to 1.5 articles/RCT in cardiovascular diseases). Compared with our primary results, our sensitivity analysis found a near identical relationship between DALYs and published trials (β=0.48, 95% CI 0.30 to 0.66; R^2^=0.13). Adjusting for multiple publications did not affect which conditions were found to have relatively high or low associated RCT publications.

In our final model, which examined the effect of the DALY location (ratio of DALYs in the top fifth of countries by socio-demographic index *vs*. the bottom four fifths), we found that DALYs, and the DALY location were both associated with increased RCT publications (β_DALY_=0.75, 95% CI 0.56 to 0.91, β_location_=0.44, 95% CI 0.38 to 0.51; adjusted R^2^ =0.33). This may be interpreted that for every 10% shift in DALYs towards high-income countries, the number of RCTs increased by 4% (3.7 to 4.9), and for every 10% increase in DALYs globally, RCTs increased by 7% (5.7 to 9.1).

## Discussion

We present the results from a natural language processing system that we used to perform a large-scale, comprehensive, automated analysis of all published RCTs indexed in the PubMed database. We find the number of published RCTs covering health conditions correlates only weakly with the burden that they inflict globally. Some conditions (particularly respiratory infections and tuberculosis, enteric infections, and transport injuries) attract substantially fewer published RCTs than expected based on the global burden.

Conditions with fewer RCTs and participants than expected tend to disproportionately affect countries with low- and medium socio-economic status. For instance, respiratory infections and tuberculosis, which had the largest disparity between global burden and number of published RCTs disproportionately affects populations in lower income countries.[24] By contrast, cancers and diabetes, for which there appears to be a relative abundance of evidence, are comparatively common in high-income countries, although this picture is changing over time.[25,26] In other words, the top fifth of countries by socio-demographic index, who conduct the vast majority of RCTs, are more closely matching RCT conduct with disease burden within their own countries (albeit still weakly). These findings suggest that a research funding strategy based on individual countries priorities does not optimally meet global needs. However, the picture is complex: global development has meant that the most important diseases affecting high-income countries (i.e. cardiovascular disease and cancers) have also become the top health priorities in low- and middle-income countries a number of years later.

The median number of participants per RCTs was 72 (i.e., a typical trial is likely to have around 36 participants per arm, or fewer trials with >2 arms). Smaller trials can be conducted faster and at lower cost, and thus can be more responsive to need, but have important limitations.[27] Small trials are more susceptible to the effects of publication bias, and have a high likelihood of generating false positive, and false negative findings.[28] Meta-epidemiological studies have shown that trials with fewer than 100 participants per arm tend to exaggerate treatment effects, compared with larger ones.[29,30] Twenty-five years after Yusuf and colleagues called for larger, simpler RCTs,[31] unfortunately we find that a majority of published trials are likely to be too small to provide useful or definitive information.

Recently published related work has used machine learning to extract sex data automatically from a large set of articles describing RCTs, finding evidence of systematic underrepresentation of women in trials.[32]Here, we broaden this approach by applying machine learning to infer the conditions and outcomes under study in trials and extracting corresponding sample sizes. By using our previously validated model[12] to automatically recognize reports of RCTs, we are able to continuously surveil the literature to maintain an up-to-date, comprehensive view of the evidence, which we make publicly available to facilitate further research.

Prior work has investigated the relationship between burden of disease and NIH funding.[8,5] These efforts relied on manual compilation of data, which necessarily limited the scope of analysis. Additionally, these efforts could only analyze conditions for which funding data was available. The authors of the landmark 1999 NEJM study noted that data was not available for conditions making up 38% of the total DALY estimate at that time.[5] Conducting this analysis manually additionally precludes the realization of a “living” view of the evidence base. By contrast, we have developed a fully automated approach that facilitates comprehensive, real-time analysis of the published RCT literature. For illustration, at the time of writing (September 2020), our system has indexed 158 RCTs examining treatments for covid-19. This contrasts with PubMed, which has indexed (largely manually) 58 with the equivalent MeSH terms.

Because our coverage of diseases is broader than prior work on the relationship between funding and burden of disease, the results are not directly comparable. However, some findings do qualitatively align with findings from prior analyses of research funding. For example, we also find that diabetes generated a relatively generous number of RCTs, as do cancers (in general). Future analyses using the methods and data could examine specific conditions within the categories presented here, investigating, for example, whether stroke is adequately researched compared with coronary artery disease, or the distribution of trials relevant to different subtypes of cancer.

Our analysis has some limitations. First, our primary analysis examines *publications* and not RCTs themselves. Our sensitivity analysis confirms previous findings that RCTs may generate more than one publication,[33] and so our estimates of publication numbers are likely to be an overestimate of the number of trials conducted. However, we found adjusting for estimates of duplicate publication made little difference to estimates of research done as a function of global burden. Second, the automated data extraction methods used here are only able to count explicit mentions of a disease or disorder, and explicit descriptions of participant numbers from the abstract, but abstracts do not always contain all the necessary information (in this study, for example, 34% of articles publications did not report the number of study participants). There is evidence of improvement over time after the publication of the CONSORT recommendations in 1996.[34]

Last, we restrict our enquiry to PubMed and therefore have not counted RCTs which have produced publications in journals not indexed in PubMed. PubMed tends to under-represent articles published in non-English languages (though includes some where an English-language translations abstracts exist). PubMed also does not index conference presentations or pharmaceutical company data repositories. By focusing on published articles, this analysis will by construction miss RCTs whose results are never published (although unpublished trial results are arguably not effectively contributing to reducing the global disease burden). Nonetheless, PubMed is reasonably comprehensive; and those RCT publications that it misses have been found to comprise a relatively small fraction of the available health research.[35,36]

Why and how do some conditions attract relatively high numbers of RCTs and trial participants? Disparities in funding may not be the sole explanation: organisational and cultural differences within medical subdisciplines could plausibly play a role. National strategies exist for increasing participation in oncology research (e.g., 2008 guidance from the American Society of Clinical Oncology which set a target of >10% of patients participating in clinical trials[37]); a large proportion of research looking at how best to increase trial recruitment has arisen from cancer RCTs.[38] Further investigation of how the better performing medical disciplines achieve their high recruitment rates might uncover strategies which are more widely applicable.

Clinical trials are one small part of tackling the global disease burden and the UN sustainable development goals, and this wider landscape needs to be considered when determining research priorities. For example, the provision of clean water is likely to be more important for preventing enteric infections than any specific health intervention, and probably does not require an RCT to prove it.[39] There may be other conditions (e.g., HIV[40]) where established treatments can enable those with the condition to have a normal lifespan without significant disability. This contrasts with some types of cancer (e.g. pancreatic cancer[41]), where we still lack reliably effective treatments. For pharmacological research, the ability to run new trials depends on the availability of new candidate drugs to test.[42] Such new candidate drugs depend upon unpredictable scientific breakthroughs, which will not neatly tally with global disease burden rankings.

Nonetheless, this data on the relative attention attracted by different health conditions may provide a useful metric to consider alongside other indicators of research need. Given that the vast majority of trials are being conducted by the wealthiest 20% of countries, targeting national health priorities is not sufficient, and a global approach is needed. We publish daily updates of the figures in this paper at www.robotreviewer.net/stateoftheevidence, and make the source code and dataset freely available via the Open Science Framework (DOI: <u>10.17605/osf.io/3db76</u>).

## Data Availability

All data are available in a public, open access repository (DOI 10.17605/osf.io/3db76).

https://osf.io/3db76/

## Contributions of authors

Initial study conceived by IJM & BCW, with intellectual input from all authors. Automatic information extraction system created by: IJM, BCW, BN, AN, FS. Curation of data for training machine learning models: JT, ANS, BCW, BN, AN, IJM. Statistical analysis designed and conducted by IJM, VL & BCW. Initial draft of manuscript by IJM & BCW. Critical revisions for important intellectual content: all; Approval of final manuscript: all

## Acknowledgements

We are enormously grateful to the volunteers from the Cochrane Crowd, who labelled hundreds of thousands of articles manually, enabling us to develop the machine learning system. Our grateful thanks to Danielle Wenner from the Department of Philosophy, Carnegie Mellon University, who reviewed early drafts and provided us with valuable feedback on the ethics of research funding distribution. We thank the Global Burden of Disease collaboration for making their data freely available for academic use.

## Data availability statement

All data are available in a public, open access repository (DOI <u>10.17605/osf.io/3db76</u>).

## Patient and Public Involvement

Patients and the public were not involved in this study.

## Footnotes

1 Model ‘training’ describes the process of adjusting model parameters to align with (typically large amounts of) data. Here, the models consume text documents (abstracts), with manual labels designating whether these are RCTs or not. Once trained (i.e., once the parameters are estimated) the model can be applied to new, unseen abstracts to provide predictions.

2 https://crowd.cochrane.org

3 MeSH refers to the Medical Subject Headings vocabulary, maintained by the National Library of Medicine (NLM) and used for indexing articles in the MEDLINE database.

